# Emergency calls are early indicators of ICU bed requirement during the COVID-19 epidemic

**DOI:** 10.1101/2020.06.02.20117499

**Authors:** By the COVID-19 APHP-Universities-INRIA-INSERM Group, Bruno Riou

**Affiliations:** Sorbonne Universite and Assistance Publique-Hopitaux de Paris, Paris, France

## Abstract

**Background:** Although the number of intensive care unit (ICU) beds is crucial during the COVID-19 epidemic caring for the most critically ill infected patients, there is no recognized early indicator to anticipate ICU bed requirements.

**Methods:** In the Ile-de-France region, from February 20 to May 5, 2020, emergency medical service (EMS) calls and the response provided (ambulances) together the percentage of positive reverse transcriptase polymerase chain reaction (RT-PCR) tests, general practitioner (GP) and emergency department (ED) visits, and hospital admissions of COVID-19 patients were recorded daily and compared to the number of COVID-19 ICU patients. Correlation curve analysis was performed to determine the best correlation coefficient (R), depending on the number of days the indicator has been shifted. A delay ≥7 days was considered as an early alert, and a delay ≥14 days a very early alert.

**Findings:** EMS calls, percentage of positive RT-PCR tests, ambulances used, ED and GP visits of COVID-19 patients were strongly associated with COVID-19 ICU patients with an anticipation delay of 23, 15, 14, 13, and 12 days respectively. Hospitalization did not anticipate ICU bed requirement.

**Interpretation:** The daily number of COVID19-related telephone calls received by the EMS and corresponding dispatch ambulances, and the proportion of positive RT-PCR tests were the earliest indicators of the number of COVID19 patients requiring ICU care during the epidemic crisis in the Ile-de-France region, rapidly followed by ED and GP visits. This information may help health authorities to anticipate a future epidemic, including a second wave of COVID19 or decide additional social measures.

**Funding:** Only institutional funding was provided.

**Research in context:** *Evidence before the study:* We searched PubMed and preprint archives for articles published up to May 17, 2020, that contained information about the anticipation of intensive care unit (ICU) bed requirement during the COVID-19 outbreak using the terms “coronavirus”, “2009-nCOV”, “COVID-19”, SARS-CoV2”, “prediction” “resource” and “intensive care”. We also reviewed relevant references in retrieved articles and the publicly available publication list of the COVID-19 living systematic review.^22^ This list contains studies on covid-19 published on PubMed and Embase through Ovid, bioRxiv, and medRxiv, and is continuously updated. Although many studies estimated the number of patients who would have severe COVID-19 requiring ICU, very few contained assessment for early signals (from internet or social media), and we retrieved no study whose data came from suspected or infected patients.

*Added values of this study:* During the COVID-19 epidemic, emergency medical system (EMS) calls, percentage of positive reverse transcriptase polymerase chain reaction (RT-PCR) tests, ambulance dispatch, emergency department (ED) and general practitioner (GP) visits of COVID-19 patients were strongly associated with COVID-19 ICU patients with an anticipation delay of 23, 15, 14, 13, and 12 days respectively. Hospitalization did not anticipated COVID-19 ICU bed requirement.

*Implication of all available evidence:* EMS calls and ambulance dispatch, percent of positive RT-PCR, and ED and GP visits could be valuable tools as daily alert signals to set up plan to face the burden of ICU bed requirement during the initial wave of the COVID-19 epidemic, and may possibly also help anticipating a second wave. These results are important since mortality has been reported being correlated to health care resources.

## Introduction

The COVID-19 pandemic has a high impact on public health in many countries.^1^ The medical response has combined all hospital resources, including emergency departments (ED), conventional hospitalization, and intensive care units (ICU). Despite the beginning of the epidemic in China in early December,^2^ most Western countries were not sufficiently prepared for its intensity and particularly the wave of critically ill patients requiring intensive care. Except for some countries which succeeded in early control of epidemic transmission chains (South Korea, Hong Kong),^3,4^ most countries (China, Italy, France, Spain, UK, USA and Brazil) experienced a rapidly diffusing epidemic pattern. It strucked the health care system with a rare violence and threatened possible ICU bed shortage which would have led to additional mortality.^5-7^ Although epidemiological analyses provided accurate early information concerning the progression of the epidemic,^8^ they were not able to predict its evolution at the peak of the crisis (including the number of ICU beds required). The peak of the crisis depends indeed on collective measures (testing, isolation of infected patients, social distancing, wearing mask, hand washing, and lockdown), which are the only actions with proven efficacy in the absence of proven specific treatment and/or vaccination to date.^9^ In France, all patients requiring ICU were admitted in such units, but this result was only obtained by a considerable increase in the number of ICU beds, and massive inter-regional ICU patient transfers, to avoid overwhelming of local ICUs.^10^

The aim of our study was to evaluate what would have been the most reliable early COVID19-related signals to anticipate ICU beds requirements. Because several days elapsed between the onset of clinical symptoms and worsening in a small proportion of infected patients requiring ICU (estimated around 5%),^11^ we hypothesized that such early signals exists and may be helpful for both public health policy or decisions and hospital management. Thus, we investigated the telephone calls received by the emergency medical services (EMS) and the immediate response provided, visits to general practitioners (GP) and emergency department (ED), hospital in-patient admissions, and positive reverse transcriptase polymerase chain reaction (RT-PCR) tests. We think that this analysis could help health care systems to more rapidly adapt to a future epidemic, including a possible second wave of COVID-19.^12^ These indicators may help health authorities to decide additional measures such as new lockdown or any other preventive measures at the population level.

## Methods

The Ile de France Region (12·1 million inhabitants) comprises 8 administrative subentities, each of them served by a medicalized EMS known as SAMU (Service d’Aide Médicale Urgente). The Paris city and its inner suburbs (6·71 million inhabitants) is covered by 4 SAMUs belonging to the Greater Paris University hospital network APHP *(Electronic supplement Figure S1)*. The individual SAMUs operate identically, use the same health information and management system (Centre d’Appel de Régulation MEdicale Nominal (CARMEN) created in 2010) and provide an adapted answer to calls to “15”, the French toll-free number dedicated to medical emergencies. This service is based on a medical response to emergency calls where an emergency physician decides the appropriate response for each case. Depending on the evaluation of the severity of the case and the circumstances, the phone response may be a medical advice, a home visit of a GP, the dispatch of an ambulance or rescue workers, and, in the most serious cases, sending a mobile intensive care unit (MICU) staffed by an emergency physician sent on scene as a second or a first tier.^13^ To cope with the surge of calls during the COVID-19 crisis, the 4 SAMUs involved in the study have increased their response capacity by creating specific procedures for COVID-19-related calls, such as staff increase, dedicated computer stations, interactive voice server, video consultation, sending instructions by SMS. Prehospital EMT and MICU teams were also significantly reinforced. Since January 20, 2020 all calls and patient records related to COVID-19 were identified in their information system and a daily automated activity report was produced.

The primary endpoint was the number of COVID19 patients who were present in ICU in the Ile de France region during the study period (from February 20 to May 5, 2020). The secondary endpoint was the daily number of new COVID19 patients admitted into ICU. During the study period, APHP staffed a regionalized and dedicated team to ensure that information concerning ICU bed availability was accurate and available in real time (Répertoire Opérationnel des Ressources Ile-de-France; https://www.ror-if.fr/ror/) and could help any physician to rapidly find an ICU bed for a given patient.^10^ We collected daily the number of ICU patients from the Système d’Information pour le Suivi des Victimes (SI-VIC) database which provides real time data on the COVID-19 patients hospitalized in French public and private hospitals (https://www.data.gouv.fr) and was activated for COVID-19 epidemic on March 13, 2020. Before that date, the number of ICU patients was collected by a direct centralized survey of the Regional Health Agency. All COVID-19 ICU cases were confirmed by RT-PCR or computed tomographic scan suggestive of SARS-CoV-2 infection.

We studied 6 indicators as they were reliable and accessible on a daily basis: 1) number of emergency calls received by the 4 SAMUs of APHP and diagnosed as suspected COVID19 patients, using the CARMEN database; 2) number of these patients requiring dispatch of an ambulance (either ordinary or MICU) using the CARMEN database; 3) number of ED visits for a clinically suspected diagnosis of COVID-19 infection in the Ile-de-France Region, using the French OSCOUR® health information system, created in 2004 and which connects all French ED (https://www.data.gouv.fr). 4) the number of COVID-19 diagnoses made by a private network of GPs who performed only emergency visits on a 24 hour and 7 day basis at home *(SOS médecins)*, in the Ile-de-France region; 5) the number of hospital admission of the COVID19 patients, in the Ile-de-France region; 6) percentage of positive RT-PCR tests for COVID19 in the Ile-de-France region. Only the percentage of positive results was considered because the availability of biological tests was markedly limited during the early phase of the epidemic. Moreover, during this early phase, only the APHP could perform RT-PCR. Publicly available sources and APHP data warehouse produced data with de-identified information.

For each indicator, we determined the onset defined as the first day the indicator became positive, 50% increase, and peak of the curve during the ascension phase. For these three points, we calculated delays as compared to endpoints. We performed correlation curve analysis during the whole study period by plotting (ICU patients at date T) *vs* (value of the indicator at date T+t) and varying t, to determine the best correlation coefficient, depending on the number of days the indicator had been shifted. The primary variable chosen to assess time delay was the correlation curve, and a time lag value ≥ 7 days was considered as an early alert indicator, and ≥ 14 days a very early one. For each indicator, we computed the time-dependant reproduction ratio (R(t)) using a gamma-distributed generation interval distribution with mean 6 days and standard deviation 4 days.^14^

We retrospectively investigated how these indicators could have been used as tools to anticipate the burden of ICU COVID19 patients. Since the initial capacity of ICU beds was 40% of that reached at the peak of the crisis we decided to fix this 40% threshold as the upper limit for each indicator, as previously reported,^15^ and half of this threshold (20%) was made the lower acceptable limit, delimiting a red zone above 40%, a green zone below 20% and an orange zone between these two limits. In addition, we also defined the slope for each indicator that correspond to the 40 and 20% of the maximum slope reached during the initial raise, using the same colour-coding.

Data are expressed as medians [interquartile IQR], or number (percentage). Correlation between two variables was assessed using the parametric Pearson test and expressed as a correlation coefficient. A P value of less than 0.05 was considered significant.

## Results

The median daily number of emergency calls received by the EMS was 1536 (IQR 494-3854), the number of emergency calls from suspected COVID19 patient ranged from 0 to 5872 (66%) The median daily number of ambulances sent was 48 (IQR 8 5-173) and the number of COVID19 patients transported to the hospital ranged from 0 to 354 (47%). The median daily number of GP visits was 91 (IQR 56-255) and the number of COVID-19 patients visited ranged from 0 to 518 (30%). The median daily number of ED visits was 519 (IQR 211-954) and the number of COVID19 ED visits ranged from 0 to 2054 (37%). The median daily number of RT-PCR was 1301 (IQR 600-3724) and the percentage of positive RT-PCR tests ranged from 0 to 54 %. The median daily number of COVID-19 hospital admission was 5552 (IQR 1148-7533), reaching a maximum of 13450. The number of ICU beds increased from 1189 to 2945 (248%) and the number of COVID ICU patients ranged from 0 to 2677 (91%). There was a high correlation between the numbers of ICU patients and new ICU COVID19 patients in the Ile-de-France region and in APHP hospitals (R^2^=0.99, P<0.0001 and R^2^=0.96, P<0.0001, respectively).

Figure 1 shows the comparison of each indicator to the primary and secondary endpoints. The number of EMS calls, number of ambulance dispatch, and percentage of positive RT-PCR tests, were very early indicators, followed by diagnosis by GP and admission to the ED. Table 1 summarizes the delay between these indicators and the primary and secondary endpoints according to the main characteristics of the curves. Correlation curve analysis is shown in *Electronic supplements Figure S2 and S3*.

**Figure 1:**
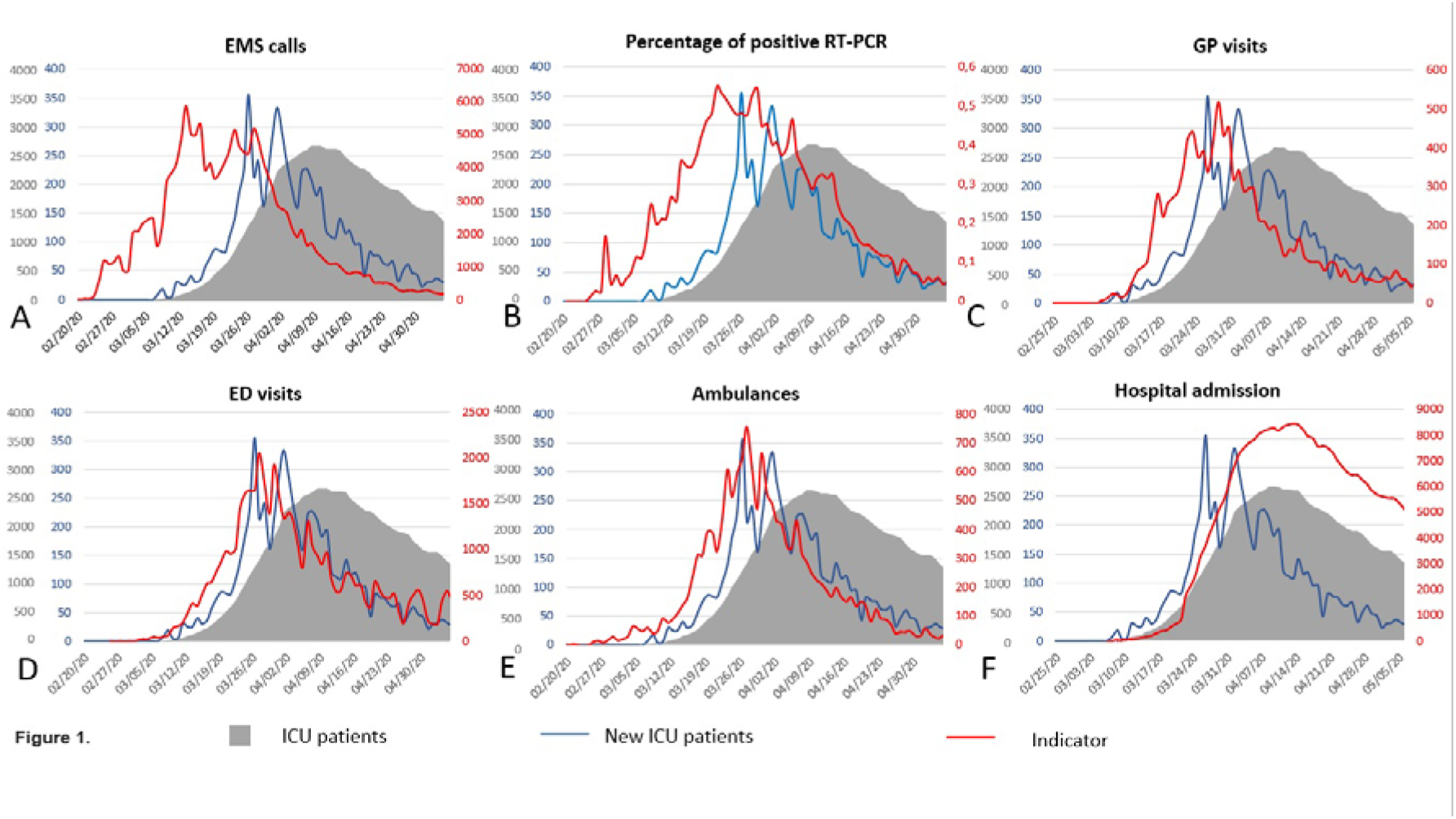
Evolution of the 6 tested indicators compared to the number of intensive care unit (ICU) patients and number of new ICU patients during the study period. EMS: emergency calls; GP: general practitioner; ED: emergency department; RT-PCR: reverse transcriptase polymerase chain reaction tests.

**Table 1.** Observed delays between each indicator and primary or secondary endpoints. Correlation curve analysis between each indicator and the number of intensive care unit (ICU) patients was performed during the whole study period (See Methods). R^2^: Pearson coefficient of correlation. EMS: emergency medical system; GP: general practitioner; ED: emergency department.

| Indicator | Onset | 50 % of the peak | Peak | Correlation curve |  |
| --- | --- | --- | --- | --- | --- |
|  | Delay<br>(Day) | Delay<br>(Day) | Delay<br>(Day) | Delay<br>(Days) | R<sup>2</sup> |
| **Primary endpoint: Number of ICU patients** |  |  |  |  |  |
| EMS calls | 16 | 18 | 26 | 23 | 0.89 |
| % of positive RT-PCR | 11 | 12 | 17 | 15 | 0.95 |
| Ambulances | 12 | 5 | 12 | 14 | 0.79 |
| ED visits | 5 | 5 | 12 | 13 | 0.84 |
| GP visits | 3 | 8 | 11 | 12 | 0.85 |
| Hospital admission | 0 | 0 | -5 | -1 | 0.99 |
| **Secondary endpoint: Number of new ICU patients** |  |  |  |  |  |
| EMS calls | 16 | 15 | 13 | 13 | 0.83 |
| % of positive RT-PCR | 11 | 9 | 4 | 4 | 0.84 |
| Ambulances | 12 | 5 | -1 | 5 | 0.87 |
| ED visits | 5 | 2 | -1 | 1 | 0.81 |
| GP visits | 3 | 5 | -2 | 4 | 0.91 |
| Hospital admission | 0 | -3 | -18 | -10 | 0.79 |

Figure 2 shows what happened if a semi-quantitative analysis of these indicators and their respective thresholds and slopes had been applied during the initial phase of the epidemic.

**Figure 2:**
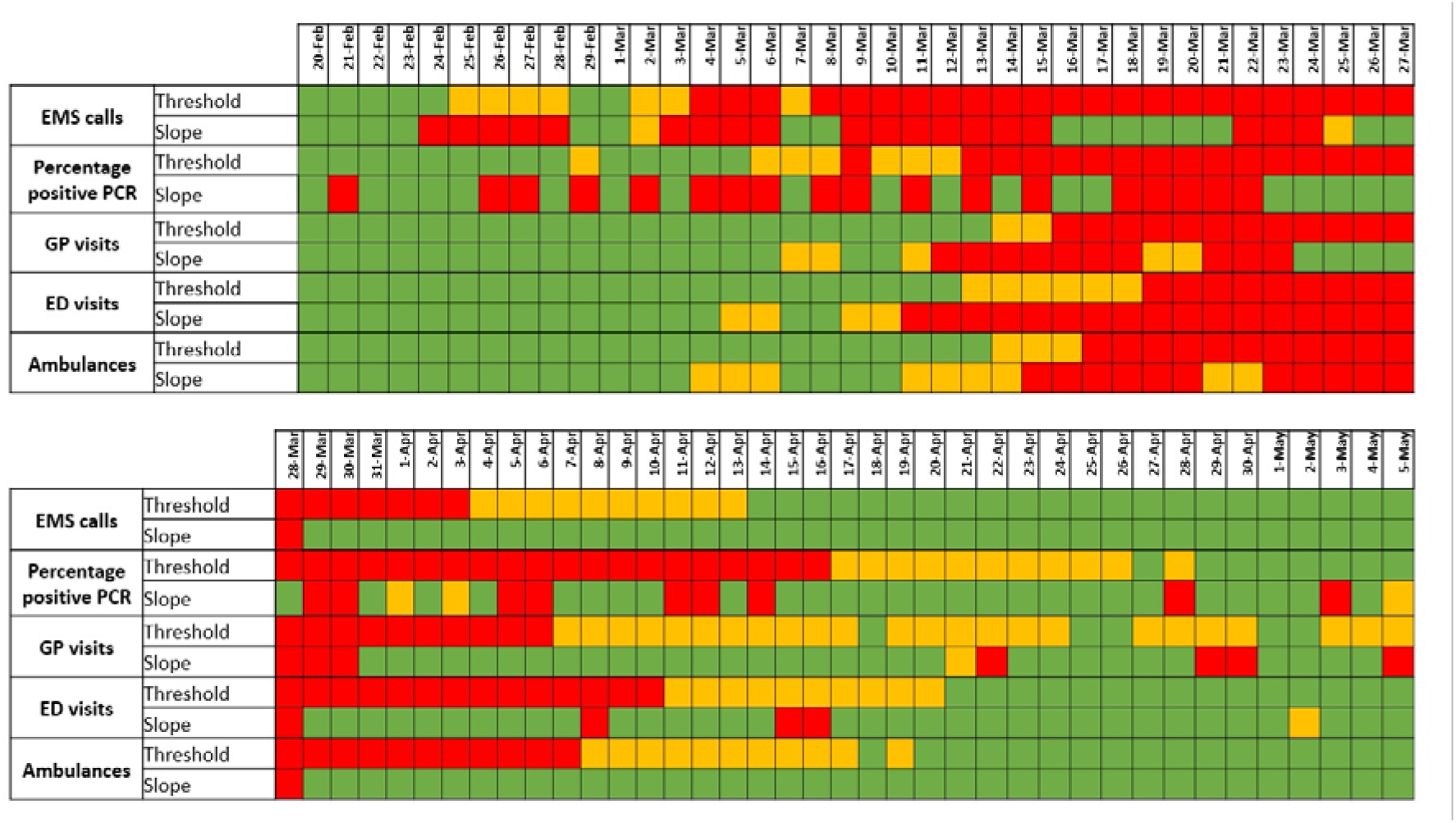
Dashboard of the retained indicators (EMS calls, percentage of positive reverse transcriptase polymerase chain (RT-PCR) tests, and general practitioners (GP) visits) and their respective thresholds and slope had been applied during the initial phase of the epidemic. Since initial capacity of ICU bed was 40% of the one reached at the peak of the crisis, a red zone was defined above this threshold, a green zone below half of this threshold (i.e. 20 % of ICU bed maximum capacity), and an orange zone between these two limits. We also defined the slope for each indicator that correspond to the 40 and 20% of the maximum slope reached during the initial raise, using the same colour coding. The first red flags would have occurred on February 24 (slope) and March 4 (threshold) for COVID19 EMS calls, 22 and 13 days before the date of the French lockdown (March 17, 2020).

The time-dependant reproduction ratio confirmed that the number of EMS calls informed early on the epidemic course (Figure 3). The effect of the lockdown on transmission was shown almost in real time, crossing the R=1 threshold 2 days after its adoption and remaining below afterwards. Similar information was obtained from other indicators with delays up to 15 days.

**Figure 3:**
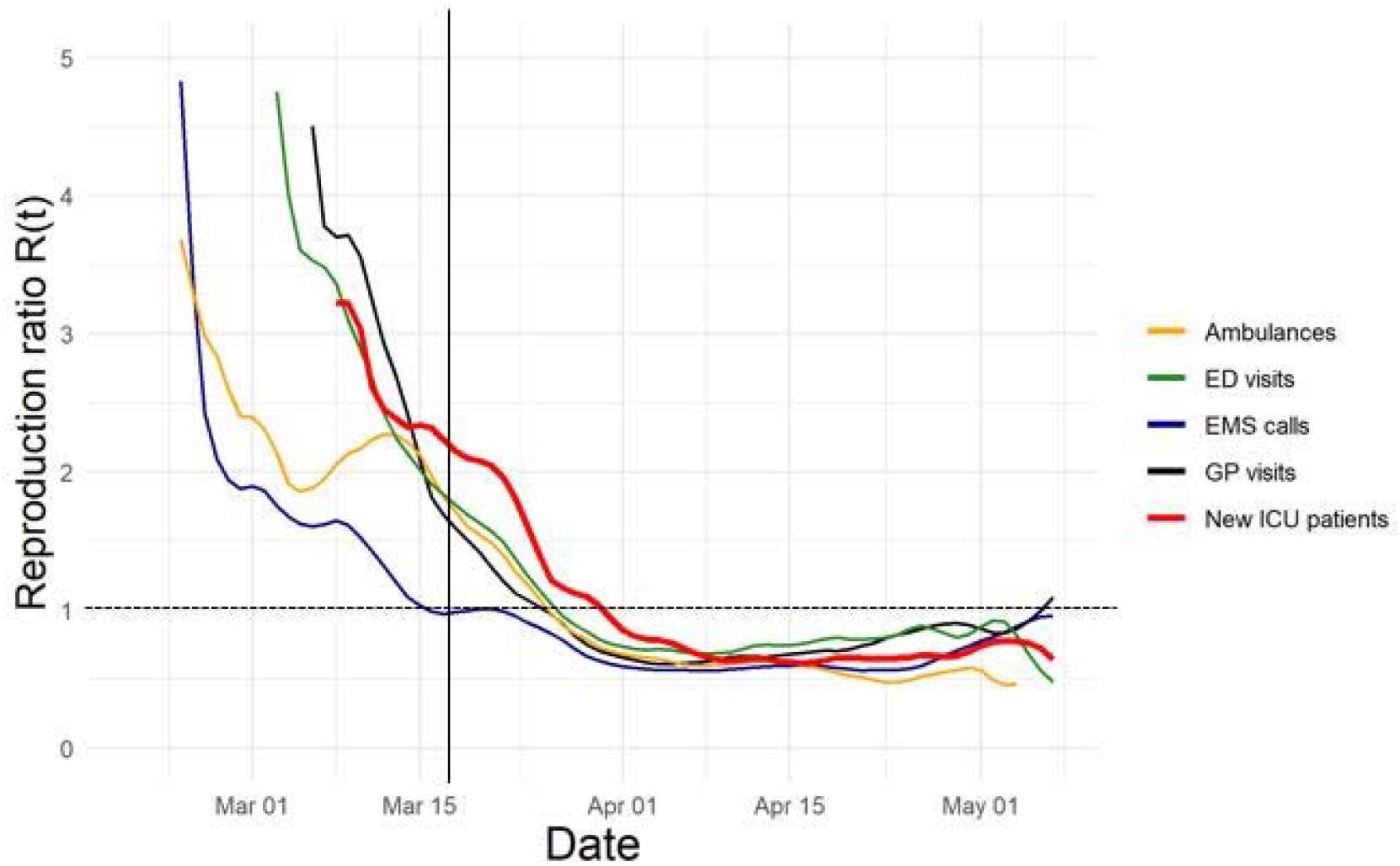
Time-dependant reproduction ratio R(t) computed from different sources of information for COVID-19: EMS calls, general practitioner (GP) visits; emergency department (ED) visits, ambulances, and compared to the number of new intensive care unit (ICU) patients. The vertical bar is the date of the lockdown in France. Positive reverse transcriptase polymerase chain test is not included in this figure because it was a percentage and not an absolute number. Hospital admission was not included because it did not anticipate number of ICU patients.

## Discussion

The present study shows that five indicators (EMS calls, percentage of positive RT-PCR tests, dispatch ambulances, ED and GP visits) anticipated the burden of ICU patients for at least 7 days during the COVID19 epidemic, the first three by at least 14 days. This result suggests that they could be valuable tools as daily alert signals to set up plan to face the outbreak burden during the initial wave of the epidemic and may possibly also work during a second wave. These results are important since mortality has been reported being correlated to health care resources.^16^

Although many studies estimated the number of patients who would have severe COVID-19,^17,18^ very few have assessed early signals associated with ICU requirements. These studies investigated internet or social media data.^19-21^ To our knowledge, no study analyzed data from COVID-19 suspected or infected patients.^22^ Since several days (estimation 7-12 days) elapse between the onset of clinical symptoms and worsening of clinical status requiring either admission to the hospital or ICU, our hypothesis is that indicators exist that could be easily available on a daily basis, enabling an accurate and anticipated survey of the COVID-19 epidemic. The availability on a defined geographical region rather than a whole country is important since a heterogeneous time and space spread of the SARS-CoV2 virus has been observed in all severely impacted countries.

Among all tested indicators, EMS calls for COVID19 was a very early one. As many countries have this type of health care organization for emergency calls, the use of this signal is widely applicable although political incitation to use this canal for the population to signal COVID19 infection may differ from one country to another. However, once appropriately used this indicator is early and sensitive. The observed delay between EMS calls and admission into ICU concords with those reported for worsening of the disease.^23^ Moreover, the medical assessment of emergency calls may be improved by learning from the first wave. The dispatch of ambulances by EMS was also a very early indicator. It should be pointed out that health authorities initially recommended symptomatic patients not to come directly to ED but rather to call the EMS which were instructed to only transport to hospitals patients needing hospital care and to refer others to GP if ambulatory care was needed. The inclusion of MICU ambulances may have also introduced a bias since some of these severe patients were directly admitted into ICU. Consequently, in different EMS systems, we cannot exclude that transport to the ED may behave differently.

The third very early indicator was the proportion of positive RT-PCR for COVID19. Many countries as well as the WHO have emphasized the importance of an early detection of the SARS-CoV2 virus by molecular diagnosis.^24^ South Korea and Germany in Europe have widely used extensive testing to better control the epidemic.^25,26^ It is therefore not surprising that this test appears as an early indicator for the COVID19 epidemic. Moreover, when testing is performed on a large scale to detect not only infected patients but also contact individuals, the signals provided by positive RT-PCR may occur earlier, which was not the case in France at the time of the first wave. The population tested further evolved with the course of the epidemic, particularly the proportion of positive patients admitted to the hospital. In France, because of initial test shortage, RT-PCR was reserved for hospitalized patients, including ICU patients, and health staff. As the epidemic diminished and RT-PCR availability increased, more tests were performed for outpatients and their respective contacts to decrease epidemic chain transmission in France. In this situation, the positive RT-PCR may become an earlier test, since some additional time elapses (estimated 3-5 days) between contamination and onset of symptoms. Further studies are required to investigate that point.

Other early indicators were the number of COVID19 diagnosis made by GP and ED. As hospitals were seen by patients as potentially dangerous, many of them were not prompted to attend the ED which led to a dramatic decrease in ED visits for any causes in France during the COVID19 epidemic, as previously reported.^28^ The use of GP diagnosis is a valuable tool if a national or regional system uploads valuable and structured information in real time. France converted an existing system for an influenza epidemic, to COVID-19 survey,^27^ but the number of involved GP remained relatively low and, as they are not available 24 hour a day, they could only enrolled a limited number of patients. The French mobile GP network SOS Médecins offers an alternative for our purpose since their members recorded appropriate clinical information concerning COVID-19 patients during the study period. This network has been included in a national process survey of epidemics since many years. Our results can be applicable to other countries when such organization is at work or when alternative and reliable GP based clinical data is collected. In addition, the French health care authorities have now promoted all GPs as key actors in the detection of COVID clusters. Therefore, evaluation and survey of GP visits should certainly become more sensitive.

Hospital admission ended up not being an early indicator of the number of ICU patients. This result concords with previous studies reporting that the delay between hospital admission to admission into ICU is closed to one day.^29^

Several limitations of this study should be noted. First, although the sample sizes were large, our observation was limited to one region of France (with a very high population density) and one event and thus extrapolation should be interpreted with caution. Second the geographical repartition and population was not identical between some indicators and endpoints *(Electronic supplement Figure S1)*. However, there was a very high correlation between ICU patients in the region and in APHP as the regulation of ICU bed availability was regionalized during the epidemic and based at the APHP. The GP indicator only reflects a particular activity (emergency visits at home) which is not distributed everywhere (less in rural areas) but this was the only accessible GP indicator. Because of biological test shortage, we only looked at the proportion of positive RT-PCR tests but the absolute number should probably be preferred in countries without such limitation. The presence of physicians (telemedicine) and not only emergency medical technicians (EMT) characterizes the French EMS model. Nevertheless, there is no indication suggesting that an EMT-based system using scripts may not lead to comparable results, particularly during an epidemic wave with a high prevalence of the disease. Definition of COVID-19 suspected diagnosis may have slightly varied during the study period in EMS, ED, and GP and between physicians. Admission into ICU has been modified during the study period as intensivists better understood the characteristics of the disease and modified their therapeutic approaches, particularly trying to avoid tracheal intubation,^30^ and transfers of ICU patients outside the region was performed just before the peak. Lastly, the raw signal of the indicators was sometimes noisy and a more advanced mathematical analysis could improve their performance.* Despite these limitations, we consider that our comparisons remain valid and could be adapted to most health systems and potentially to other types of epidemic scheme.

## Conclusion

The daily number of COVID19-related telephone calls received by the EMS and corresponding ambulance dispatch, and the proportion of positive RT-PCR were the earliest indicators of the number of COVID19 patients requiring ICU care during the epidemic crisis in the Ile-de-France region, rapidly followed by ED and GP visits. This information may help health authorities to anticipate a future epidemic, including a second wave of COVID19, to monitor lockdown exit and decide additional social measures to better control COVID-19 outbreak.

^*^Gaubert S and colleagues. Forecasting the local progression of the COVID-19 epidemic from medical emergency calls. https://arxiv.org/abs/2005.14186

## Data Availability

Some data were extracted from a publicly available source others from Assistance Publique-Hopitaux de Paris (Paris, France) warehouse (Entrepot des Donnees de Sante, EDS) are available on request.

## Acknowledgment

We thank Dr. David Baker, DM, FRCA, (Department of Anesthesiology and Critical Care, Hôpital Necker-Enfants Malades, Paris, France) for reviewing the manuscript. The authors are deeply grateful to Audrey Bourdette, Arthur Cornet, Alban Jourdain, Côme Cheritel, and Henri Matalon from the Data science team, Department of Strategy and transformation at APHP (Paris, France) for their essential assistance for data collection, analysis, editing of figures and statistical analysis. We also thank Pr. Laurent Tréluyer (Department of informatics and computer sciences, APHP, Paris, France) for providing us data and Dr. Christophe Leroy (APHP, Paris, France) for valuable discussion.

## Conflict of interest

All authors have completed and submitted the ICMJE Form for Disclosure of Potential Conflicts of Interest. No disclosure was reported.

## Authors’ contributions

Batteux and Riou had full access to all the data in the study and takes responsibility for their integrity and the accuracy. *Study concept and design:* Batteux, Carli, Gaubert, and Riou. *Acquisition and verification of the data:* Adnet, Batteux, Calvez, Carli, Chansard, Hausfater, Lecarpentier, Loeb, Paugam, Riou, and Vieillard-Baron. *Drafting of the manuscript:* Batteux and Riou. *Critical revision of the manuscript for important intellectual content:* all authors. *Statistical analysis:* Batteux and Boëlle. *Study supervision:* Batteux, Carli, and Riou

## Fundings

Support was provided only by institutional sources

## Transparency statement

The corresponding author affirms that this manuscript is an honest, accurate and transparent account of the study being reported; that no important aspects of the study have been omitted; and that any discrepancies from the study as planned have been explained.

## Data sharing

Some data were extracted from a publicly available source (https://www.data.gouv.fr) others from APHP warehouse (Entrepôt des Données de Santé, EDS) are available on request to Pr. Batteux.

**Electronic Supplement Figure S1.**
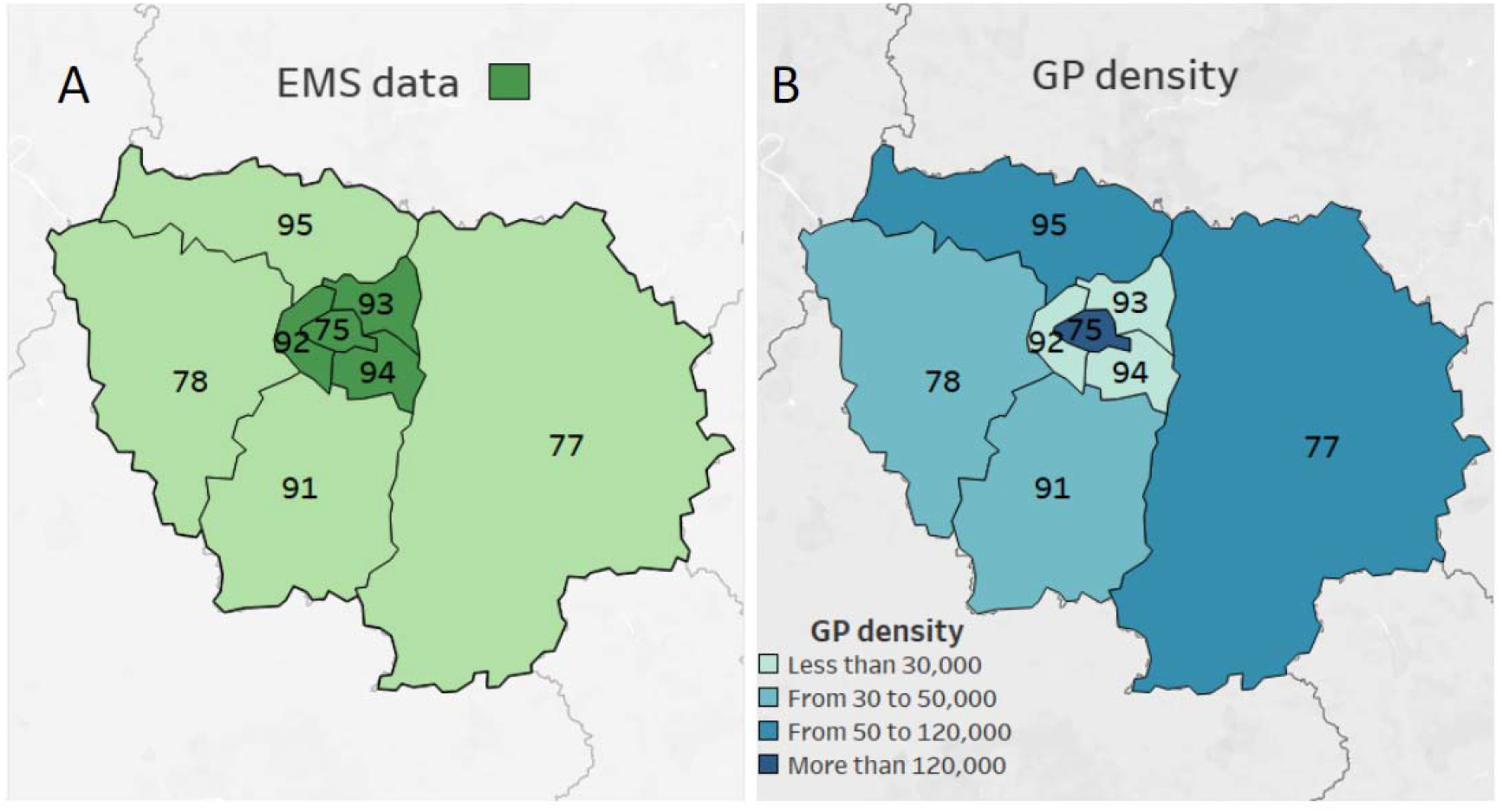
The Ile-de-France region (12·1 million inhabitants) comprises 8 administrative sub-identities, indicated by their number on the map, the town of Paris being 75. In the present study, the numbers of emergency departments (ED) visits, positive reverse transcriptase polymerase chain reaction (RT-PCR) tests, hospital admissions, intensive care unit (ICU) patients, and new ICU patients were obtained from the whole region. A regionalized organization was installed enabling to rapidly find an ICU bed for a given patient wherever the patient was initially admitted. Panel A: Data from emergency medical system (EMS), including emergency calls and dispatch of ambulances were obtained from the Paris city (75) and its inner suburbs which comprise 4 administrative sub-entities (75, 92, 93, 94) and their respective EMS (6·71 million inhabitants). Panel B: Data from general practitioner (GP, SOS Médecins network) were obtained from the Ile-de-France region but the density of activity of this GP network (expressed as number of annual visits per million inhabitants).is heterogeneous within the Ile-de-France region.

**Electronic Supplement Figure S2.**
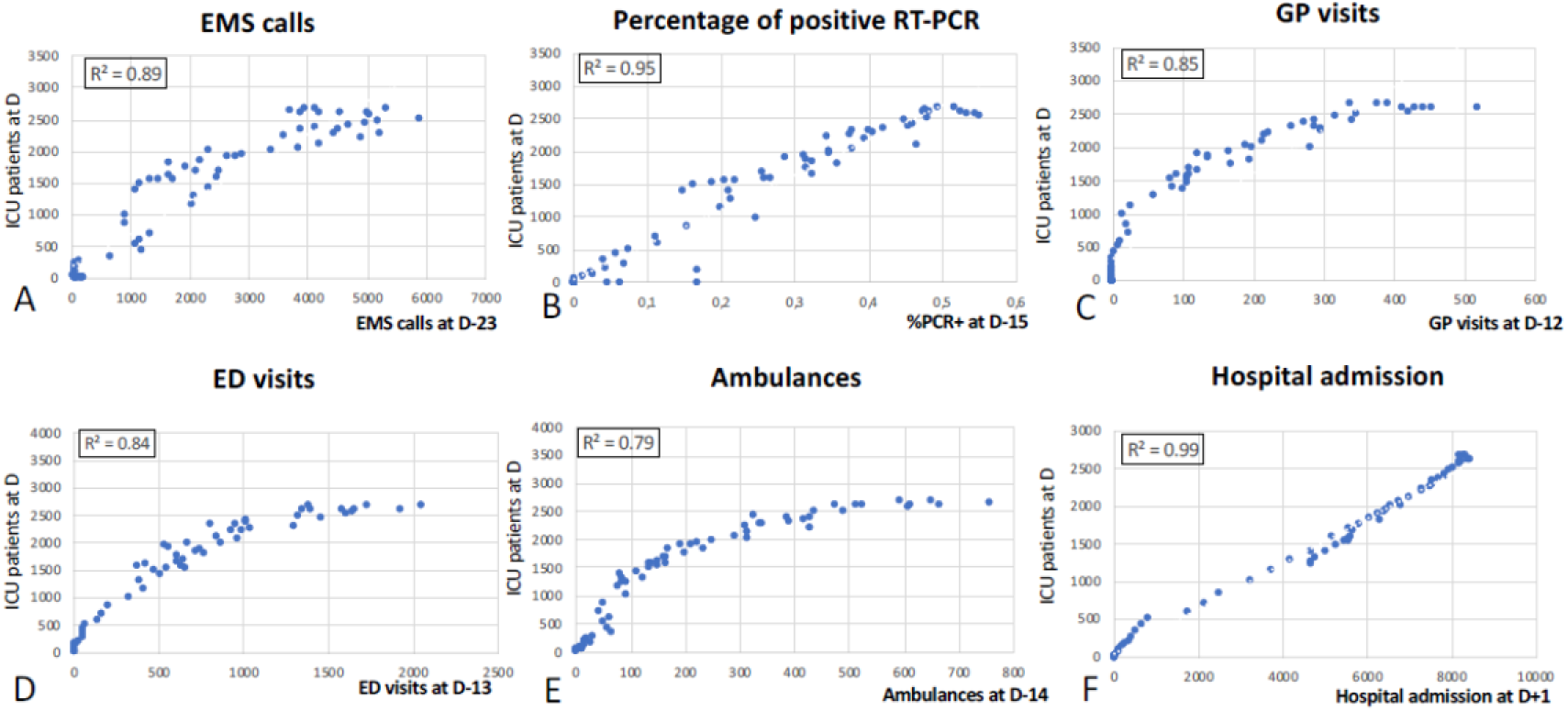
Correlation curves of the 6 tested indicators compared to the number of intensive care unit (ICU) patients during the study period. EMS: emergency calls; GP: general practitioner; ED: emergency department. D: delay (in days) between the two variables. R^2^: Pearson coefficient of correlation.

**Electronic Supplement Figure S3.**
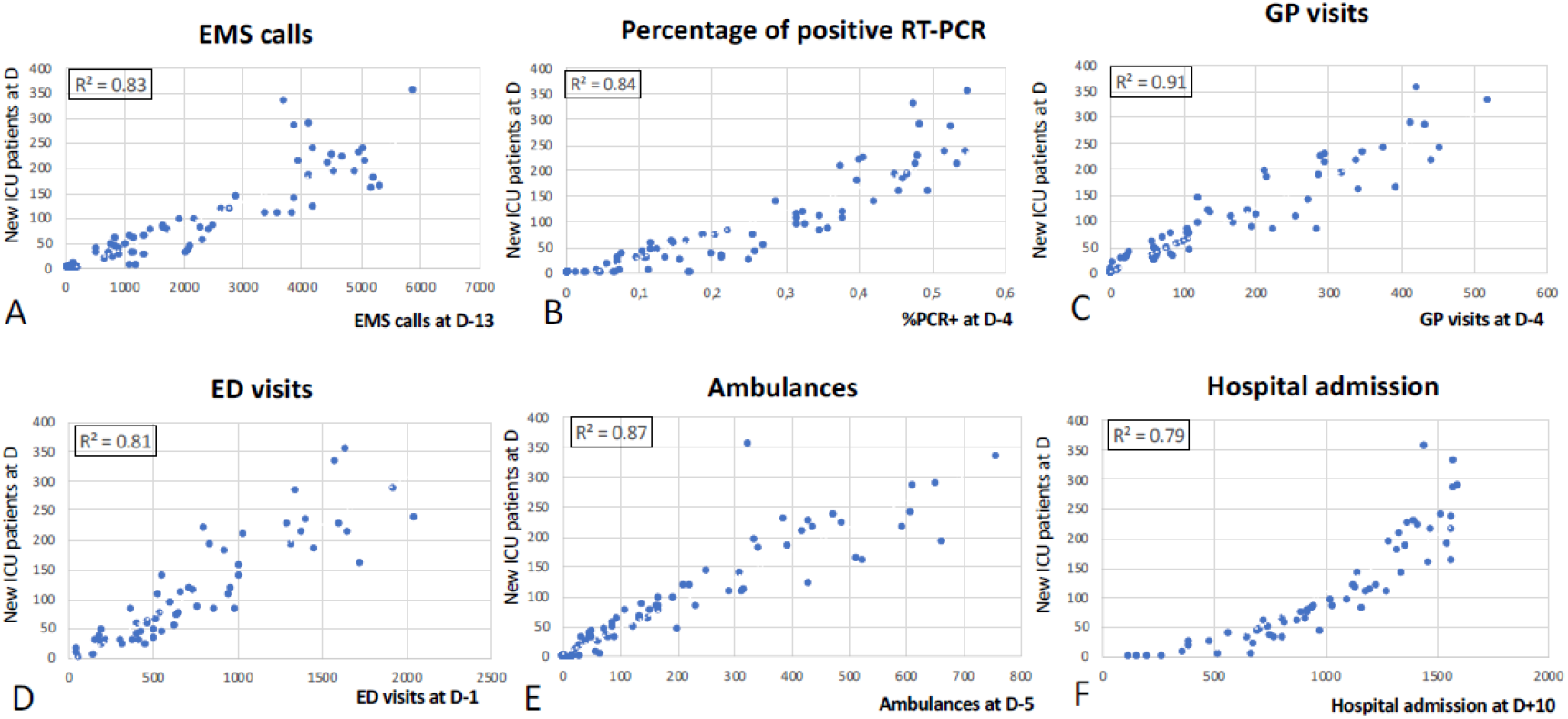
Correlation curves of the 6 tested indicators compared to the number of new intensive care unit (ICU) patients during the study period. EMS: emergency calls; GP: general practitioner; ED: emergency department. D: delay (in days) between the two variables. R^2^: Pearson coefficient of correlation.

## Notes

### Competing Interest Statement

The authors have declared no competing interest.

### Clinical Trial

Study performed on anomymised data publicly available

### Author Declarations

This study was approved by the Sorbonne Universite ethical committee (CER Number 2020-55, Paris, France). Patient consent was not applicable.

